# A modular pipeline for natural language processing-screened human abstraction of a pragmatic trial outcome from electronic health records

**DOI:** 10.1101/2025.06.23.25330134

**Authors:** Robert Y. Lee, Kevin S. Li, James Sibley, Trevor Cohen, William B. Lober, Janaki O’Brien, Nicole LeDuc, Kasey Mallon Andrews, Anna Ungar, Jessica Walsh, Elizabeth L. Nielsen, Danae G. Dotolo, Erin K. Kross

**Author notes:** **Address correspondence to:** Robert Y. Lee, MD, MS, Assistant Professor of Medicine, Division of Pulmonary, Critical Care, and Sleep Medicine, Cambia Palliative Care Center of Excellence at UW Medicine, University of Washington School of Medicine, Seattle, WA, Address: Harborview Medical Center, 325 9th Ave., Box 359762, Seattle, WA 98104.

## Abstract

**Background:** Natural language processing (NLP) allows efficient extraction of clinical variables and outcomes from electronic health records (EHR). However, measuring pragmatic clinical trial outcomes may demand accuracy that exceeds NLP performance. Combining NLP with human adjudication can address this gap, yet few software solutions support such workflows. We developed a modular, scalable system for NLP-screened human abstraction to measure the primary outcomes of two clinical trials.

**Methods:** In two clinical trials of hospitalized patients with serious illness, a deep-learning NLP model screened EHR passages for documented goals-of-care discussions. Screen-positive passages were referred for human adjudication using a REDCap-based system to measure the trial outcomes. Dynamic pooling of passages using structured query language (SQL) within the REDCap database reduced unnecessary abstraction while ensuring data completeness.

**Results:** In the first trial (N=2,512), NLP identified 22,187 screen-positive passages (0.8%) from 2.6 million EHR passages. Human reviewers adjudicated 7,494 passages over 34.3 abstractor-hours to measure the cumulative incidence and time to first documented goals-of-care discussion for all patients with 92.6% patient-level sensitivity. In the second trial (N=617), NLP identified 8,952 screen-positive passages (1.6%) from 559,596 passages at a threshold with near-100% sensitivity. Human reviewers adjudicated 3,509 passages over 27.9 abstractor-hours to measure the same outcome for all patients.

**Conclusion:** We present the design and source code for a scalable and efficient pipeline for measuring complex EHR-derived outcomes using NLP-screened human abstraction. This implementation is adaptable to diverse research needs, and its modular pipeline represents a practical middle ground between custom software and commercial platforms.

## Background

Advances in natural language processing (NLP) are creating new opportunities for pragmatic clinical trialists to measure important outcomes from free-text electronic health records (EHR). To date, much work in clinical NLP has focused on tasks well-suited to existing methods such as discrete data element extraction (*e*.*g*., diagnoses or symptoms), named entity recognition (*e*.*g*., persons, organizations, or locations), and text classification (*e*.*g*., assigning sentiment or topical categories).^1,2^ However, using NLP to measure pragmatic clinical trial outcomes from the EHR presents two key challenges. First, many patient-centered outcomes—such as symptoms, quality of life, adverse events, and functional outcomes—are documented in clinical language that is both linguistically complex and also highly sensitive to context, making them challenging to measure accurately. Second, the high cost and rigor of clinical trials demands low tolerance for measurement error and misclassification, which are inherent risks of NLP-based approaches. For these reasons, hybrid measurement strategies that combine NLP with final adjudication of outcomes by humans will likely remain essential for clinical trialists. Yet, software solutions are scarce for such hybrid NLP pipelines, which may lead investigators to either forgo such approaches entirely or turn to homegrown software and databases.

Our research group has conducted several randomized clinical trials examining the effect of communication-priming interventions on the primary outcome of EHR-documented goals-of-care discussions,^3–5^ an important clinician-documented process measure in palliative care research. In many health systems, such discussions are documented in unstructured text and can be difficult and costly to extract from voluminous EHRs. Furthermore, the outcome of goals-of-care discussions is linguistically and conceptually complex,^6–8^ requiring extensive training of human abstractors to measure consistently. To facilitate measurement of this outcome at scale, our group has developed multiple NLP models to identify documented goals-of-care discussions. Many of these models have shown promising performance, yet remain insufficiently accurate for stand-alone use in measuring the primary outcome of a clinical trial.^9–12^

Our group sought to measure EHR-documented goals-of-care discussions in two clinical trials evaluating the efficacy of communication-priming interventions designed to promote goals-of-care discussions and their documentation for hospitalized patients with serious illness. To do this, we implemented a two-step outcome measurement process combining an NLP screening instrument with human adjudication. We used a previously-described deep-learning NLP model to screen EHR passages,^11^ then referred screen-positive passages for human adjudication in a process designed to maximize abstraction efficiency. We implemented a pipeline for NLP-screened human abstraction using common research software and data platforms such as REDCap (version 12.2.1; Vanderbilt University, project-redcap.org) and Stata (version 18.0; StataCorp, stata.com).^13^

In this report, we describe and provide the source code for this NLP-screened human abstraction pipeline. We also discuss design parameters and pragmatic and statistical implications that may be applicable to other studies. We specifically focus on the design and implementation of the human adjudication pipeline, which exists separately from the screening NLP model^11^ and is portable to different screening modalities and outcomes.

## Methods

### Design and Implementation

#### Problem description, and NLP screening of EHR passages

In the United States, healthcare norms often default to providing life-extending treatments to all patients.^14^ However, some patients with chronic life-limiting illness prefer a palliative or comfort-focused approach that emphasizes symptom relief over life extension.^15–17^ To better align treatments with patients’ values, goals and preferences, experts have highlighted the need for high-quality communication between patients, surrogate decision-makers, and clinicians when making decisions about life-sustaining treatments.^18,19^ These conversations, commonly called “goals-of-care discussions,” play an important role in delivering goal-concordant care.^6,19–22^ Given the frequent handoffs and transitions in modern healthcare, experts also agree on the importance of documenting goals-of-care discussions in the EHR and communicating them across the continuum of care.^19,23–25^ Documented goals-of-care discussions are an important process measure and research outcome in the field of serious illness communication.^19,26^ However, despite their importance, documented goals-of-care discussions represent a small proportion of EHR text, and are rarely represented as structured data. Additionally, the construct of a goals-of-care discussion is linguistically and medically complex, encompassing multiple dimensions and existing along continua rather than as discrete entities (**Figure 1**).^10,27^ These factors make the process of measuring documented goals-of-care discussions through conventional chart abstraction both costly and labor-intensive; and, training human abstractors is further challenged by the inconsistent and infrequent nature of such documentation in the EHR.^9,28^

**Figure 1.**
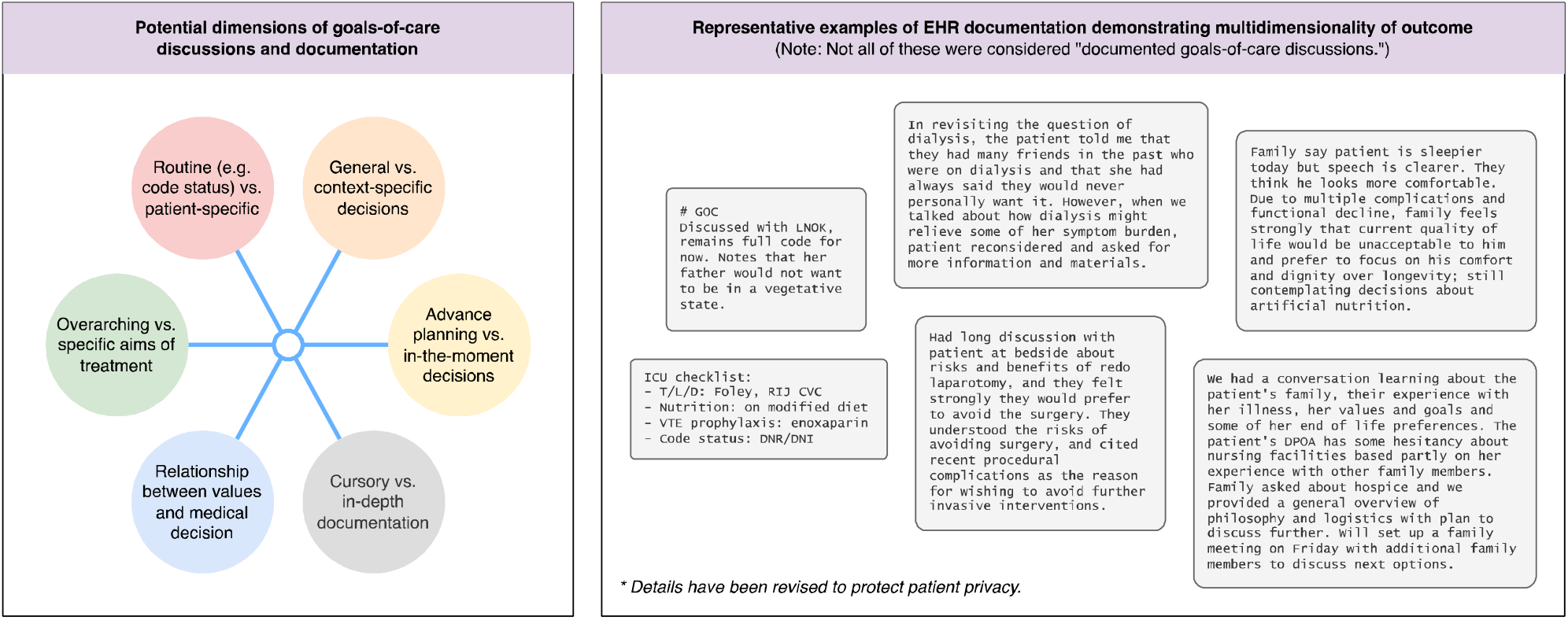
Dimensions and examples of EHR documentation related to goals of care. Abbreviations: GOC, goals of care; TLD: tubes, lines, drains; RIJ CVC: right internal jugular central venous catheter; VTE: venous thromboembolism; DNR/DNI: do not resuscitate / intubate; DPOA: durable power of attorney.

In this report, we describe the NLP-screened human abstraction pipeline for two trials in the Project to Improve Communication about Serious Illness (PICSI) series: PICSI Hospital Trial 1 (hereafter, “Trial 1”; ClinicalTrials.gov ID NCT04281784), a 2,512-patient pragmatic trial conducted April 2020 – March 2021;^4,5^ and, PICSI Hospital Trial 2 (hereafter, “Trial 2”; ClinicalTrials.gov ID NCT04283994), a 617-patient comparative effectiveness trial conducted July 2021 – November 2023.^4^ Both trials evaluated the effect of communication-priming interventions on the primary outcome of EHR-documented goals-of-care discussions as measured by NLP-screened human abstraction. The NLP screening instrument used for both trials was a fine-tuned instance of the freely available BioClinicalBERT^29,30^ deep learning model (BERT: Bidirectional Encoder Representations from Transformers)^31^ that was trained on a manually-labeled corpus of 4,642 clinical notes from the PICSI Pilot Trial (hereafter, “Training dataset”), a 150-patient pilot trial examining the same outcome and conducted November 2018 – February 2020.^3^ Notes were tokenized (*i*.*e*., split into pre-defined words and sub-words, called tokens) using the BioClinicalBERT dictionary and further divided into non-overlapping passages (hereafter, “BERT segments”) of ≤ 512 tokens each (the upper limit for BERT’s sequence length) based on common whitespace patterns. Manual labeling of the training dataset was performed by a team of investigators and research staff using a previously-published codebook^11 Supplement 1, eAppendix 2^ and a qualitative data analysis platform (Dedoose, Sociocultural Research Consultants, dedoose.com), with regular meetings and co-review of coded passages to reconcile coding and ensure consistency. The training dataset was labeled over a total of 287 abstractor-hours during a 4-month period.^11^

Characteristics of the EHR corpora are presented in **Table 1**. The volume of EHR text within which documented goals-of-care discussions could be found was very large in both trials, with Trial 1 yielding 44,324 eligible notes, and Trial 2 yielding 11,574 eligible notes. Moreover, documented goals-of-care discussions represented only 0.2% of EHR passages (BERT segments) in the training dataset. Because the standalone BERT NLP classifier did not reach sufficient levels of precision and recall for the study, trial investigators elected to measure the primary outcome using an NLP-screened human abstraction pipeline.^11^

**Table 1.** Characteristics of electronic health record note corpora.

|  | Training dataset | Trial 1 dataset | Trial 2 dataset |
| --- | --- | --- | --- |
| No. patients | 150 | 2,512 | 617 |
| No. (%) patients with GOCD | 34 (23) | – | – |
| No. notes per patient, median (IQR) | 19 (11-29) | 12 (7-22) | 14 (9-23) |
| No. EHR notes | 4,642 | 44,324 | 11,574 |
| No. (%) notes with GOCD | 340 (7) | – | – |
| Note length, words, median (IQR) | 1070 (1578-3202) | 1039 (688-1468) | 983 (622-1695) |
| No. BERT segments ($\leq 512$ tokens) | 201,394 | 2,644,387 | 559,596 |
| No. (%) BERT segments with GOCD | 446 (0.2) | – | – |

Abbreviations: GOCD, [documented] goals-of-care discussion; EHR, electronic health record; IQR, interquartile range; BERT, Bidirectional Encoder Representations from Transformers.

#### Configuring REDCap for human adjudication of screened passages

Although many software packages exist for annotating unstructured text to support NLP training, there are fewer solutions available for the rapid adjudication of text for categorical outcomes. This task requires streamlined, efficient tools that minimize interactional overhead for rapid and scalable workflows. The most widely cited software for this use, ClinicalRegEx (Lindvall Lab, Dana-Farber Cancer Institute),^32,33^ is constrained to regular expression-based screening. Other low-overhead solutions such as Local Turk (Dan Vanderkam)^34^ lack support for multiple abstractors or remote access. Custom-built end-user databases (*e*.*g*., Microsoft Access) offer tailored functionality but require substantial effort to configure and rarely meet modern standards for multi-user performance, web accessibility, reliability or security.

Many clinical researchers are familiar with REDCap,^13^ a web-accessible platform designed for secure, multi-user data collection in research settings. Although not originally intended for unstructured text review, REDCap’s flexible form builder supports adaptation to a wide range of research workflows, and its application programming interfaces (APIs) allow for automated dataset management. We developed a REDCap instrument for efficient human adjudication of large numbers of NLP-screened text passages. To import EHR data into REDCap, we encoded screen-positive NLP passages into comma-separated values (CSV) files containing candidate EHR passages, their surrounding context, and pertinent metadata, creating one REDCap record per candidate passage. We also incorporated features to flag passages requiring co-review. Detailed REDCap configurations and other design considerations are provided in **Appendix A**; our configuration is also available from the REDCap Shared Library (access instructions in **Appendix A**).^35,36^ Stata source code for preparing segmentized EHR data for contextual display in REDCap is provided in **Appendix B**.

#### Targeted human adjudication via dynamic record-pooling

The primary outcome of the trial was the occurrence of a documented goals-of-care discussion at any time within 30 days of randomization. To inform potential future analyses, the investigators also sought to measure days between randomization and *first* documented goals-of-care discussion. Abstractors were blinded to randomization and NLP-predicted probabilities, and we used open-source rule-based NLP software to redact patient, clinician, and family names from passages.^37^

Preliminary data from the training dataset indicated that goals-of-care documentation often appears multiple times in a single patient’s record, either due to copy-pasted text or multiple distinct discussions. To minimize unnecessary abstraction, we aimed to limit human adjudication to EHR passages occurring between randomization and the earliest *human-confirmed* documented goals-of-care discussion for each patient, or through the end of the outcome assessment period if no such documentation was found. This created a unique challenge: the set of passages requiring abstraction for the study outcome could not be determined in advance, as it depended on the outcome of the abstraction itself. To address this, we developed a dynamic record-pooling process that ran in parallel with ongoing abstraction, removing passages occurring after a confirmed goals-of-care discussion for the same patient from the abstraction dataset (**Figure 2**). Additionally, the efficiency of record-pooling also depended on the sequence in which records were abstracted. To reduce unnecessary abstraction while avoiding a sequential, per-patient approach (*e*.*g*., abstracting all records for patient 1, then patient 2, and so on)—which could introduce adverse incentives to code for positive outcomes—we shuffled screen-positive EHR passages across patients while preserving temporal ordering within each patient. This produced a random-appearing sequence of passages that remained optimized for dynamic record-pooling efficiency (**Figure 3**; Stata source code in **Appendix C**).

**Figure 2.**
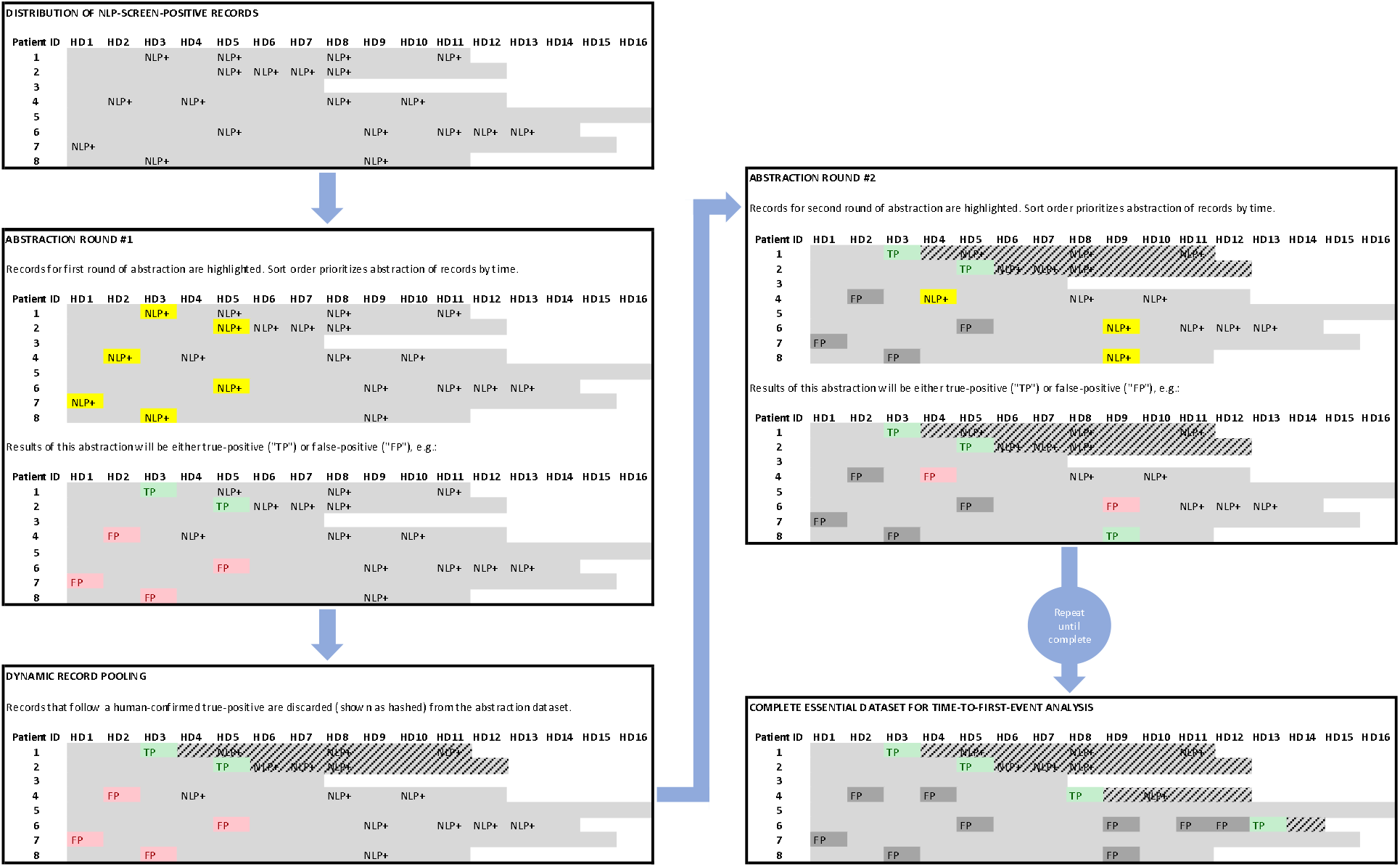
Abstraction process for achieving minimum complete dataset for time-to-first-event data with dynamic record-pooling. Data provided are hypothetical, and have been simplified to reflect one electronic health record (EHR) passage per note per hospital day (HD). For this depiction, abstraction has been divided into hypothetical “rounds” of one record per patient, partitioned by dynamic record-pooling events. “NLP+” denotes screen-positive records. Results of human adjudication of screened records are denoted as “TP” (true-positive) or “FP” (false-positive). Records discarded by dynamic record-pooling are shown as hashed. Actual sequence of passages adjudicated in each “round” of abstraction may vary by sort order (see **Figure 3**), and actual dynamic record-pooling may take place at any time (see **Appendix D**), *i*.*e*., not necessarily between “rounds.”

**Figure 3.**
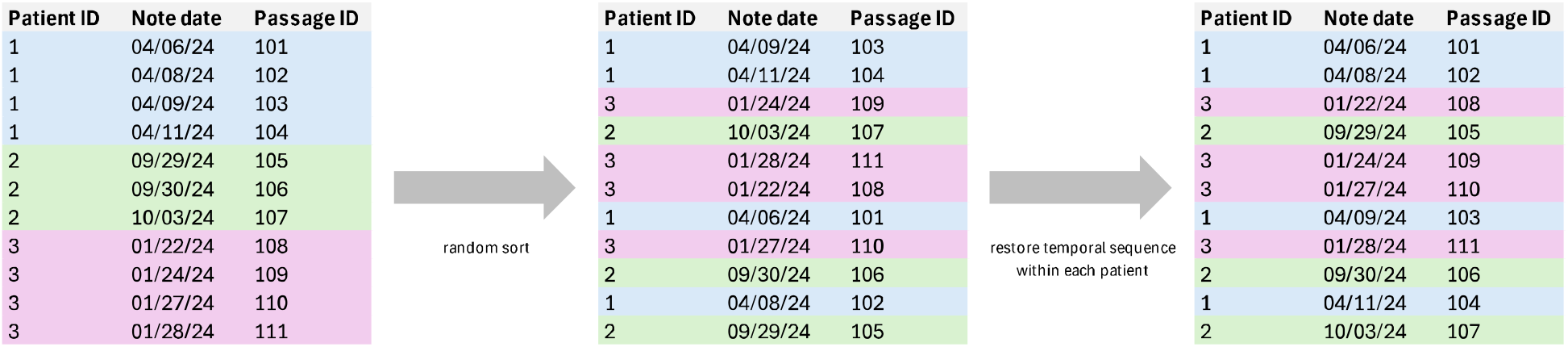
Temporal-sequence-preserving sort for human adjudication of time-to-event data.

A key challenge of dynamic record-pooling is that very few data abstraction platforms have functionality to alter the abstraction dataset in real-time. Owing to the large number of EHR passages in our project, it was impractical to pool records manually. To modify the abstraction dataset at regular intervals, we implemented an automated script that retrieved a deidentified report of human-adjudicated outcomes from the REDCap database, then used this data to determine which unabstracted passages needed to be removed from, or replaced back into, the abstraction dataset. The script was built to be robust to changes in data (*e*.*g*., handling of changing abstraction results following co-review), and leveraged REDCap’s sorting behaviors to non-destructively discard or replace records from the abstraction dataset without deleting or re-creating REDCap records. **Appendix D** provides the source code for a Bourne-again shell (bash v3.2) script implementing dynamic record-pooling using structured query language (SQL) for a REDCap database for collecting time-to-first-event data.

#### Quality control of human adjudications

Abstractors underwent a training curriculum and group abstraction work to achieve consistency in applying the abstraction codebook. All human adjudications were subject to multiple quality-control measures, including regular meetings to discuss sampled human-adjudicated passages, functionality for abstractors to “flag” specific passages for group discussion, periodic audits of individual abstractors’ work, and in select instances, referral to clinician-investigators for additional review to arrive at a final adjudication.

### Statistical Implications

The statistical implications of any two-stage outcome measurement process such as NLP-screened human abstraction must be carefully considered in any study. In our study, we assumed that the specificity of an NLP-screened human-abstracted outcome would be 100% relative to a manually-abstracted gold standard. However, the sensitivity of the outcome measure is dependent on both the performance of the NLP screening instrument and selection of the screening threshold. Misclassification from an imperfect NLP screening instrument can affect study power and findings of association. We have previously reported on the power implications of non-differential outcome misclassification in randomized trials that use NLP-screened human abstraction with imperfect sensitivity.^11^

In considering NLP-screened human abstraction of patient-level outcomes, in which records are screened and adjudicated at a fine level of measurement (*i*.*e*., the EHR passage) and patient outcomes are defined by aggregation of passage-level measures, it is important to note two things: (1) that performance characteristics at passage- and patient-levels are likely to differ substantially, in ways that are dependent on the nature of the aggregation step between passage- and patient-level outcomes; and, (2) that the patient-level sensitivity of the NLP instrument at a given threshold *k* is <u>not</u> equal to the patient-level sensitivity of NLP-screened human abstraction at the same threshold *k*. This second observation may seem counterintuitive, but it is explained by the fact that the posterior testing step in NLP-screened human abstraction only refers screen-positive passages for human adjudication. As a result, some patients who would have true-positive patient-level outcomes when measured by an NLP-only instrument would have false-negative patient-level outcomes when measured by NLP-screened human abstraction at the same discriminating threshold, because the NLP instrument may both fail to detect any true-positive passage-level outcomes (thereby withholding positive passage-level outcomes from the human abstractor) while also detecting false-positive passages that are then appropriately adjudicated as negative by the human abstractor. These observations lead to the conclusions that (1) estimating patient-level performance from validation data requires either validation data that is collected at the patient level, or advanced statistical methods that may require additional assumptions for validity; and, (2) estimating patient-level sensitivity of NLP-screened human abstraction similarly requires validation data measured at both the passage and patient level.

## Results

### Trial Enrollment and Text Corpora

Over April 2020 to March 2021, Trial 1 enrolled 2,512 patients that were randomized into two arms: 1,255 to a pragmatic clinician-facing-only communication-priming intervention (Jumpstart Guide), and 1,257 to usual care. Over July 2021 to November 2023, Trial 2 enrolled 617 patients that were randomized into three arms: 203 to a bilateral patient- and clinician-facing Jumpstart Guide, 205 to a clinician-facing-only Jumpstart Guide, and 209 to usual care with enrollment surveys. The total number of eligible EHR notes for the primary outcome was 44,324 in Trial 1, and 11,574 in Trial 2 (**Table 1**).

### NLP Validation and Screening Threshold Selection

Nearing conclusion of enrollment for Trial 1, trial investigators and the data and safety monitoring committee requested an interim analysis with two objectives: (1) to validate patient- and note-level performance of the NLP screening instrument in the trial population, and (2) to estimate the prevalence of the primary outcome by manual chart abstraction in a sample of trial participants enrolled to date, with emphasis on the prespecified subgroup of patients with Alzheimer disease and related dementias (ADRD) due to an enrollment shortfall in this group.^4,5^ In February 2021, we randomly sampled 160 patients (80 with ADRD) from all participants who had been randomized at least 30 days prior. We then assembled all eligible notes from these patients’ charts into a validation corpus. One patient without ADRD was mistakenly linked to the wrong encounter and excluded, resulting in a final sample of 159 patients, 80 (50%) with ADRD, who contributed 2,480 eligible notes.

To measure the primary outcome prevalence, a separate team of investigators and research staff (excluding trial principal investigators, and distinct from those who abstracted the training dataset) performed conventional whole-chart abstraction of the validation corpus for documented goals-of-care discussions. Abstraction procedures and quality control were the same as those used for the training dataset. Manual abstraction identified 268/2,480 (11%) notes from 54/159 (34%) patients containing documented goals-of-care discussions.^11^ Prevalence of goals-of-care discussions was similar between patients with and without ADRD (25/80 [31%] vs. 29/79 [37%], p=0.51).

Following completion of manual abstraction, we evaluated the performance of the NLP screening instrument in the validation sample using receiver operating characteristic curve analysis and precision-recall analysis. We examined both the performance of the NLP classifier *alone*, as well as the expected performance of *NLP-screened human abstraction* through conditional analysis of the validation sample across candidate NLP screening thresholds. The methods and results of these evaluations have been previously reported.^11^ Based on the results of this evaluation, we selected a screening threshold of 0.5 (an uncalibrated BERT score, range [0,1]), which corresponded to NLP passage-level sensitivity of 83.7%, specificity of 99.2%, and precision of 35.9%; note-level sensitivity of 96.2%, specificity of 77.9%, and precision of 37.6%; and, patient-level sensitivity of 99.2%, specificity of 33.2%, and precision of 45.6%. The patient-level sensitivity of NLP-screened human abstraction at this threshold was 92.6% (binomial 95CI 87.6%, 97.6%).^11^

### NLP-Screened Human Abstraction of Trial Outcomes

At the selected NLP screening threshold, the BioClinicalBERT classifier identified 22,187 of 2.6 million (0.8%) EHR passages representing 11,287 of 44,324 (25%) notes and 1,957 of 2,512 patients (80%) to refer for human adjudication. Of the 22,187 passages referred for human adjudication, dynamic pooling allowed us to adjudicate only 7,494 (33.8%) passages to achieve complete time-to-first-event data. These 7,494 passages were adjudicated by a team of 3 abstractors over 34.3 abstractor-hours in a 3-week period. For comparison, conventional chart abstraction was estimated to require approximately 2,000 abstractor-hours for this dataset.^11^ In a random sample of 300 NLP-positive EHR passages adjudicated by multiple abstractors, inter-abstractor agreement was 89.7% (Fleiss’ kappa 0.65).

For Trial 2, investigators were able to select a lower (more sensitive) screening threshold due to the smaller number of participants and eligible records. To maximize sensitivity, investigators selected a screening threshold corresponding to the 98.5^th^ percentile BERT score (an uncalibrated constant of 0.00279598). The selected threshold was 100% sensitive for documented goals-of-care discussions in the Trial 1 validation sample; validation data from Trial 2 is still undergoing collection. Using this screening threshold, the BioClinicalBERT classifier identified 8,952 of 559,596 (1.6%) EHR passages representing 3,718 of 11,574 (32%) notes and 533 of 617 (86%) patients to refer for human adjudication. Of these 8,952 passages, 3,509 (39%) were adjudicated by a team of 8 abstractors over 27.9 abstractor-hours in a 2-month period to achieve complete time-to-first-event data. For comparison, conventional chart abstraction was estimated to require approximately 500 abstractor-hours for this dataset.

## Discussion

By combining common research software and data platforms such as REDCap and Stata with our research group’s separately-developed BERT-based NLP infrastructure, we were able to implement a streamlined and efficient workflow for large-scale NLP-screened human abstraction of a complex linguistic outcome. Our pipeline leveraged REDCap APIs for automated dynamic record-pooling to achieve complete data with maximal efficiency. Although our implementation is not an “out-of-the-box” solution for all clinical researchers, it provides a starting point for measurement of EHR outcomes via two-step measurement strategies such as automated screening and subsequent human adjudication. Our approach is portable to different NLP screening methods and models, as they are implemented upstream of the human adjudication pipeline; and, it may be further adapted to other types of measures and different essential datasets with nominal programming effort. By implementing this pipeline in a modular fashion using common software and platforms, our approach also maximizes flexibility for future deployments in other projects while maintaining a low-friction user experience and minimizing maintenance costs. Using secure platforms such as REDCap ensures a baseline level of data security often overseen at an institutional level, while still allowing simultaneous abstraction efforts by multiple individuals over a web-based platform.

In studies where the event of interest is rare, dynamic record-pooling as described here can significantly reduce the burden of human abstraction. In Trial 1, there were 22,187 screen-positive passages at the selected threshold; human adjudication of all screen-positive passages would have required over 100 abstractor-hours. By dynamically discarding passages following a human-confirmed goals-of-care discussion for the same patient, we were able to achieve complete data for the measures of interest (*i*.*e*., cumulative incidence and time-to-first-event) abstracting only 7,494 (33.8%) of the referred passages over 34.3 abstractor-hours—a 66% reduction.^11^ Investigators considering dynamic record-pooling must carefully consider the essential dataset of EHR passages for their type of measure, and how best to prioritize or discard passages for human adjudication to achieve data completion with maximal efficiency and minimal bias. In **Table 2**, we discuss process considerations for implementing NLP-screened human abstraction of three common types of measures—time-to-event, cumulative incidence, and incidence rate—alongside general considerations that apply to all measures.

**Table 2.**
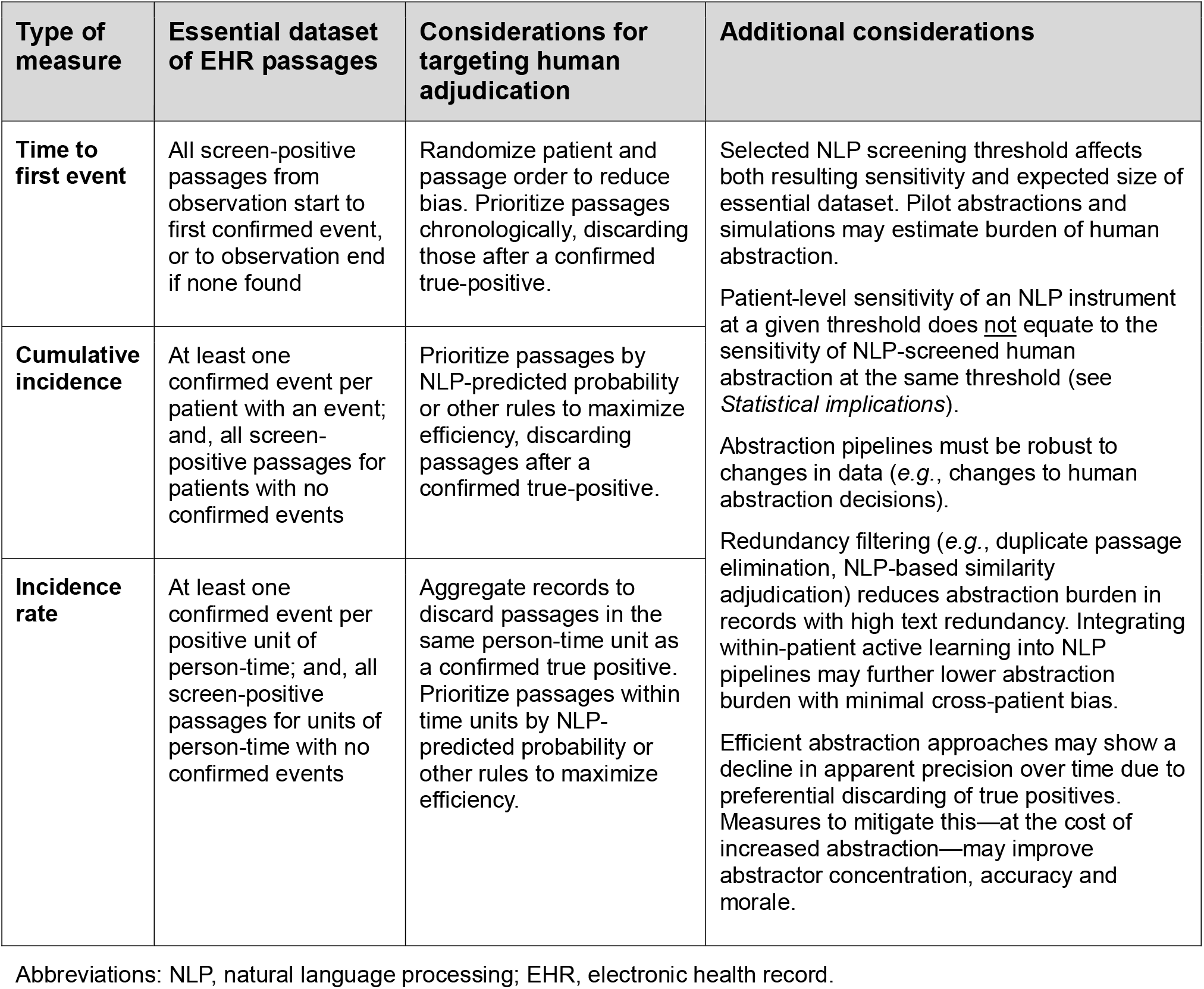
Process considerations for NLP-screened human abstraction of rare events.

In our use case, NLP performance was evaluated at the note- and patient-level for binary outcomes (ever vs. never).^11^ During NLP-screened human abstraction efforts, time-to-first-event data were collected only to support potential future analyses. However, for time-to-event analyses, NLP-screened human abstraction could also bias results if the first occurrence of the outcome is not correctly identified. Researchers applying this approach to time-to-event studies should explicitly assess NLP performance in capturing the earliest positive passage per patient.

Despite the advantages of such two-stage measurement approaches, adopting these approaches to measure complex outcomes also introduces important limitations to consider. First, the robustness of an NLP-screened approach to outcome measurement is heavily dependent on the robustness of the screening NLP instrument. In addition to loss of sensitivity, biases in the instrument are likely to propagate into the resulting measurements. As NLP models become increasingly “black box”-like, it becomes of paramount importance for investigators to critically evaluate the model performance across potential confounding variables. Second, biases inherent in pragmatically collected EHR data may also threaten both internal and external validity. For example, variability in EHR documentation across different clinical settings may lead to loss of construct validity, confounding, or loss of generalizability. Third, although the programming burden of implementing such a pipeline is not high by programmers’ standards, it still requires substantial technical expertise and access to the requisite platforms and servers. REDCap—relied on heavily in our implementation—is often unavailable outside of academic medical centers, limiting the scalability of our implementation. Fourth, improper configuration may expose study participants to data security risks.

As EHR data become an increasingly common source of pragmatic measures in clinical research, investigators must become familiar with the many ways one may extract EHR data and choose collection processes wisely. Close collaboration between clinical researchers, informaticists, and front-line research staff is important to ensuring fidelity to the clinical construct of interest, minimizing bias, promoting efficiency while maximizing accuracy, and ensuring that all data is handled securely.

## Conclusion

To measure a linguistically complex EHR-documented outcome in two randomized trials, we combined previous work in NLP with common research software and data platforms to construct a pipeline for collecting NLP-screened human-adjudicated outcome measures using a streamlined and efficient web-based user interface with dynamic record-pooling to minimize abstraction time. The methodological principles, source code and data instrument design are easily adaptable to other research projects that use EHR-screened human abstraction or any other two-stage measurement strategy requiring human adjudication. Our implementation represents an appealing middle-ground between custom software solutions and repurposed commercial products for researchers working with EHR data.

## Data Availability

Due to the identifiable nature of the protected health information evaluated in this study, freetext data are not available for disclosure without institutional approval and authorization.

## Author contributions

Dr. Lee had full access to all of the data in the study and takes responsibility for the integrity of the data and the accuracy of the data analysis.

## Acknowledgement

The authors are grateful for the contributions of the late J. Randall Curtis, MD, MPH, who was a founding principal investigator of this research program.

