## Appendix A for "A modular pipeline for natural language processing-screened human abstraction of a pragmatic trial outcome from electronic health records"

### Appendix A. REDCap configuration for human adjudication of NLP-screened EHR passages.

REDCap versions 10.x-14.x instrument definition for human adjudication of natural language processing (NLP)-screened electronic health record (EHR) passages. Instrument definition is provided in JavaScript Object Notation (JSON) format for readability. An importable version of the instrument is available from the REDCap Shared Library located at [https://redcap.vumc.org/consortium/library/search.php](https://redcap.vumc.org/consortium/library/search.php) (instrument title: "NLP-screened EHR passage adjudication", uploaded January 22, 2025); access requires session authentication via a local REDCap project in Development or Draft Mode, under Project Setup > Design your data collection instruments > Explore the REDCap Instrument Library.

**Notes:**

- `record_id` refers to the REDCap identifier for the EHR passage. By default, records are sorted in order of `record_id` in the REDCap interface; and, when a user clicks "Save & Go To Next Record" at the end of a form, REDCap automatically sends the user to the next `record_id` by sort order. As such, users wishing to specify the order in which passages are adjudicated should sort `record_id` in the desired order of abstraction. Furthermore, application programming interfaces (APIs) may leverage this behavior to discard or replace passages from the minimum complete dataset at regular intervals without deleting or re-uploading records.
- `pid`, `note_eventid`, and `segment_id` are patient, EHR note, and EHR passage identifiers, respectively. The patient identifier is used by API calls to discard passages from the minimum complete dataset. Not all of these fields will be useful for all use cases. For security purposes, hashing patient-identifying values of these identifiers is recommended to minimize protected health information (PHI) available to API calls.
- `segment_txt` contains pre-processed EHR passage text *and* adjoining passages (denoted and formatted by `<span>` blocks specifying alternate formatting). Adjoining passages, when present, should be handled by abstractors in accordance with project-specific guidelines. `segment_txt_nocontext` contains EHR passage text without adjoining passages, which may be useful for reports.
- Variables beginning with `display_` are used to customize the instrument user interface for optimal efficiency and accuracy. REDCap versions 12.x-14.x respect the cascading style sheet (CSS) `style` attribute of `div` and `span` Hypertext Markup Language (HTML) tags, which allows for custom formatting beyond that facilitated by REDCap's interactive interface.
- Abstractors were instructed to "bite off what they can chew" when selecting a beginning record ID for abstraction. The provision of `display_notice` prompted abstractors when their progress "collided" with another abstractor's efforts. Alternative worflows may include pre-assigning records to abstractors, or dividing records into groups that abstractors may self-assign; however, such methods may require additional adjustments in how records are dynamically discarded or replaced in response to abstraction results.


```json
[
  {
    "Variable / Field Name": "record_id",
    "Field Type": "text",
```

```json
    "Field Label": "Record ID",
    "Choices, Calculations, OR Slider Labels": null,
    "Field Note": null,
    "Text Validation Type OR Show Slider Number": null,
    "Identifier?": null,
    "Branching Logic (Show field only if...)": null,
    "Required Field?": null,
    "Custom Alignment": null,
    "Field Annotation": null
  },
  {
    "Variable / Field Name": "abstractor_user",
    "Field Type": "text",
    "Field Label": "Abstractor ID",
    "Choices, Calculations, OR Slider Labels": null,
    "Field Note": "Auto-populated for first abstraction.",
    "Text Validation Type OR Show Slider Number": null,
    "Identifier?": null,
    "Branching Logic (Show field only if...)": null,
    "Required Field?": true,
    "Custom Alignment": null,
    "Field Annotation": " @USERNAME"
  },
  {
    "Variable / Field Name": "abstractor_datetime",
    "Field Type": "text",
    "Field Label": "Abstraction Date/Time",
    "Choices, Calculations, OR Slider Labels": null,
    "Field Note": "Auto-populated for first abstraction.",
    "Text Validation Type OR Show Slider Number": "datetime_seconds_mdy",
    "Identifier?": null,
    "Branching Logic (Show field only if...)": null,
    "Required Field?": true,
    "Custom Alignment": null,
    "Field Annotation": " @NOW"
  },
  {
    "Variable / Field Name": "display_notice",
    "Field Type": "descriptive",
    "Field Label": "<div style=\"background-color:#ff0000;color:#ffffff;text-align:center;\"><span style=\"font-size:72pt;\">STOP.</span>\n<span style=\"font-size:16pt;\">This record has already been previously abstracted.\nPlease make sure you intend to review or modify this record.</span></div>",
    "Choices, Calculations, OR Slider Labels": null,
    "Field Note": null,
    "Text Validation Type OR Show Slider Number": null,
    "Identifier?": null,
    "Branching Logic (Show field only if...)": null,
    "Required Field?": null,
```

```
    "Custom Alignment": null,
    "Field Annotation": " @IF([outcome_human]='' AND [abstractor_user]!='TheDupinator_OriginalTBD',@HIDDEN,'')"
  },
  {
    "Variable / Field Name": "pid",
    "Field Type": "text",
    "Field Label": "Imported: pid (hashed)",
    "Choices, Calculations, OR Slider Labels": null,
    "Field Note": null,
    "Text Validation Type OR Show Slider Number": null,
    "Identifier?": null,
    "Branching Logic (Show field only if...)": null,
    "Required Field?": true,
    "Custom Alignment": null,
    "Field Annotation": " @HIDDEN"
  },
  {
    "Variable / Field Name": "note_eventid",
    "Field Type": "text",
    "Field Label": "Imported: event_id (hashed)",
    "Choices, Calculations, OR Slider Labels": null,
    "Field Note": null,
    "Text Validation Type OR Show Slider Number": null,
    "Identifier?": null,
    "Branching Logic (Show field only if...)": null,
    "Required Field?": true,
    "Custom Alignment": null,
    "Field Annotation": " @HIDDEN"
  },
  {
    "Variable / Field Name": "segment_id",
    "Field Type": "text",
    "Field Label": "Imported: Segment ID",
    "Choices, Calculations, OR Slider Labels": null,
    "Field Note": null,
    "Text Validation Type OR Show Slider Number": null,
    "Identifier?": null,
    "Branching Logic (Show field only if...)": null,
    "Required Field?": true,
    "Custom Alignment": null,
    "Field Annotation": " @HIDDEN"
  },
  {
    "Variable / Field Name": "segment_txt",
    "Field Type": "notes",
    "Field Label": "Imported: Segment text",
    "Choices, Calculations, OR Slider Labels": null,
    "Field Note": null,
```

```
    "Text Validation Type OR Show Slider Number": null,
    "Identifier?": true,
    "Branching Logic (Show field only if...)": null,
    "Required Field?": true,
    "Custom Alignment": "LH",
    "Field Annotation": " @READONLY @HIDDEN"
  },
  {
    "Variable / Field Name": "segment_txt_nocontext",
    "Field Type": "notes",
    "Field Label": "Imported: Segment text, without context",
    "Choices, Calculations, OR Slider Labels": null,
    "Field Note": null,
    "Text Validation Type OR Show Slider Number": null,
    "Identifier?": true,
    "Branching Logic (Show field only if...)": null,
    "Required Field?": true,
    "Custom Alignment": "LH",
    "Field Annotation": " @READONLY @HIDDEN"
  },
  {
    "Variable / Field Name": "display_segment",
    "Field Type": "descriptive",
    "Field Label": "Segment text:\n\n<div style=\"background-color:#e3eaf9;font-weight:normal;padding:.75em;font-size:12pt;\">[segment_txt]</div>\n<span style=\"font-weight:normal;font-family:Courier,Courier New;font-size:8pt;\">pid-hashed [pid], <span style=\"color:#cc0000;\">eid-hashed [note_eventid]</span>, sid [segment_id]</span>",
    "Choices, Calculations, OR Slider Labels": null,
    "Field Note": null,
    "Text Validation Type OR Show Slider Number": null,
    "Identifier?": null,
    "Branching Logic (Show field only if...)": null,
    "Required Field?": null,
    "Custom Alignment": null,
    "Field Annotation": null
  },
  {
    "Variable / Field Name": "outcome_human",
    "Field Type": "radio",
    "Field Label": " \nPRIMARY OUTCOME: Does the text shown above represent a goals-of-care discussion, or discussion of new ACP/DPOA?\n\n<span style=\"font-weight:normal;\">[Reference: <a href=\"#\" target=_blank>Link to Codebook</a>. We are ONLY interested in text meeting criteria for codes GOCD or ACP/DPOA.]</span>\n ",
    "Choices, Calculations, OR Slider Labels": "1, YES - GOCD or ACP/DPOA | 0, NO - does not meet criteria for either code",
```

```
    "Field Note": "<br>For text that is SUBTHRESHOLD, JUST CODE STATUS, LIVING IN THE PAST, CONGRESS, etc., please respond \"NO\" to this question.<br><br>Surrounding context (grayed text) is provided IN CASE it is helpful, but reading it is optional. That being said, IF you happen to notice that the grayed-out text meets criteria for GOCD or ACP/DPOA, please respond \"YES\" to this question.",
    "Text Validation Type OR Show Slider Number": null,
    "Identifier?": null,
    "Branching Logic (Show field only if...)": null,
    "Required Field?": true,
    "Custom Alignment": "LV",
    "Field Annotation": null
  },
  {
    "Variable / Field Name": "display_verdict_blank",
    "Field Type": "descriptive",
    "Field Label": "<div style=\"font-size:48pt;color:#ffffff;text-align:center;\"> </div>",
    "Choices, Calculations, OR Slider Labels": null,
    "Field Note": null,
    "Text Validation Type OR Show Slider Number": null,
    "Identifier?": null,
    "Branching Logic (Show field only if...)": "isblankormissingcode([outcome_human])",
    "Required Field?": null,
    "Custom Alignment": null,
    "Field Annotation": null
  },
  {
    "Variable / Field Name": "display_verdict_goc",
    "Field Type": "descriptive",
    "Field Label": "<div style=\"font-size:48pt;color:#ffffff;text-align:center;background-color:#00cc00;\">+GOC!</div>",
    "Choices, Calculations, OR Slider Labels": null,
    "Field Note": null,
    "Text Validation Type OR Show Slider Number": null,
    "Identifier?": null,
    "Branching Logic (Show field only if...)": "[outcome_human]=1",
    "Required Field?": null,
    "Custom Alignment": null,
    "Field Annotation": null
  },
  {
    "Variable / Field Name": "display_verdict_negative",
    "Field Type": "descriptive",
    "Field Label": "<div style=\"font-size:48pt;color:#ffffff;text-align:center;background-color:#cc0000;font-weight:normal;\">Negative</div>",
    "Choices, Calculations, OR Slider Labels": null,
    "Field Note": null,
    "Text Validation Type OR Show Slider Number": null,
```

```json
    "Identifier?": null,
    "Branching Logic (Show field only if...)": "!isblankormissingcode([outcome_human]) AND [outcome_human]=0",
    "Required Field?": null,
    "Custom Alignment": null,
    "Field Annotation": null
  },
  {
    "Variable / Field Name": "abstractor_comments",
    "Field Type": "notes",
    "Field Label": "Abstractor comments (memos)",
    "Choices, Calculations, OR Slider Labels": null,
    "Field Note": "NOTE: Comments are only for memos and tracking. They are NOT analyzed by the investigators. If you have questions or concerns about a particular segment, please flag the segment for review.",
    "Text Validation Type OR Show Slider Number": null,
    "Identifier?": true,
    "Branching Logic (Show field only if...)": null,
    "Required Field?": null,
    "Custom Alignment": "LH",
    "Field Annotation": null
  },
  {
    "Variable / Field Name": "abstractor_flag",
    "Field Type": "checkbox",
    "Field Label": "Abstractor flags",
    "Choices, Calculations, OR Slider Labels": "1, Flag this segment for further review | 2, This segment was CO-REVIEWED",
    "Field Note": null,
    "Text Validation Type OR Show Slider Number": null,
    "Identifier?": null,
    "Branching Logic (Show field only if...)": null,
    "Required Field?": null,
    "Custom Alignment": "LV",
    "Field Annotation": null
  },
  {
    "Variable / Field Name": "abstractor_user2",
    "Field Type": "text",
    "Field Label": "Co-reviewer ID(s)",
    "Choices, Calculations, OR Slider Labels": null,
    "Field Note": null,
    "Text Validation Type OR Show Slider Number": null,
    "Identifier?": null,
    "Branching Logic (Show field only if...)": "[abstractor_flag(2)]",
    "Required Field?": null,
    "Custom Alignment": null,
    "Field Annotation": null
  },
```

```
  {
    "Variable / Field Name": "abstractor_datetime2",
    "Field Type": "text",
    "Field Label": "Co-review Date/Time",
    "Choices, Calculations, OR Slider Labels": null,
    "Field Note": null,
    "Text Validation Type OR Show Slider Number": "datetime_seconds_mdy",
    "Identifier?": null,
    "Branching Logic (Show field only if...)": "[abstractor_flag(2)]",
    "Required Field?": null,
    "Custom Alignment": null,
    "Field Annotation": null
  },
  {
    "Variable / Field Name": "coreview_outcome",
    "Field Type": "dropdown",
    "Field Label": "Co-review Reason & Outcome",
    "Choices, Calculations, OR Slider Labels": "1, FLAGGED RECORD - co-reviewed to follow-up and resolve flag | 2, RANDOM co-review; I AGREED with the previous abstractor's decision | 3, RANDOM co-review; I REVERSED the previous abstractor's decision | 4, Other reason for co-review; I AGREED with the previous abstractor's decision | 5, Other reason for co-review; I REVERSED the previous abstractor's decision | 99, DO NOT USE - For flagging rule-based auto-daemons only",
    "Field Note": null,
    "Text Validation Type OR Show Slider Number": null,
    "Identifier?": null,
    "Branching Logic (Show field only if...)": "[abstractor_flag(2)]",
    "Required Field?": null,
    "Custom Alignment": "LH",
    "Field Annotation": null
  },
  {
    "Variable / Field Name": "display_coreviewrem2",
    "Field Type": "descriptive",
    "Field Label": "<div style=\"background-color:#cc0000;font-size:14pt;color:#ffffff;\"> <br>REMINDER: <span style=\"font-weight:normal;\">Please uncheck the abstractor's flag if you are resolving it.<br> </span></div>",
    "Choices, Calculations, OR Slider Labels": null,
    "Field Note": null,
    "Text Validation Type OR Show Slider Number": null,
    "Identifier?": null,
    "Branching Logic (Show field only if...)": "[coreview_outcome]=1 AND [abstractor_flag(1)]",
    "Required Field?": null,
    "Custom Alignment": null,
    "Field Annotation": null
  },
  {
    "Variable / Field Name": "display_coreviewrem",
```

```
    "Field Type": "descriptive",
    "Field Label": "<div style=\"color:#cc0000;background-color:#ffff00;font-size:14pt;\">NOTE: Please make sure you have changed the previous abstractor's response for the PRIMARY OUTCOME question <span style=\"font-weight:normal;\">(\"Does the text shown above represent...\")</span> to the correct and final response.</div>",
    "Choices, Calculations, OR Slider Labels": null,
    "Field Note": null,
    "Text Validation Type OR Show Slider Number": null,
    "Identifier?": null,
    "Branching Logic (Show field only if...)": "[coreview_outcome]!=\"\"",
    "Required Field?": null,
    "Custom Alignment": null,
    "Field Annotation": null
  },
  {
    "Variable / Field Name": "display_gofast",
    "Field Type": "descriptive",
    "Field Label": "Abstractor workflow note:\n<div style=\"align:center;font-size:14pt;font-weight:normal;\">For this project, we will track record statuses using abstraction results and flags. You may <span style=\"background-color:#ffff00;\">skip</span> the \"Complete?\" drop-down menu below, and select \"Save & Go To Next Record\" from the blue drop-down button to proceed.</div>",
    "Choices, Calculations, OR Slider Labels": null,
    "Field Note": null,
    "Text Validation Type OR Show Slider Number": null,
    "Identifier?": null,
    "Branching Logic (Show field only if...)": null,
    "Required Field?": null,
    "Custom Alignment": null,
    "Field Annotation": null
  }
]
```


#### Additional configuration options

##### Custom REDCap record label

To streamline abstraction, we applied a custom record label to our project so that each record's automatically-generated abstraction status would be visible on all Record Status Dashboards without requiring abstractors to manually change the record status field. In REDCap version 14, this setting is located under Project Setup > Additional Customizations > Set a Custom Record Label. We specified a custom record label of:

```
Record abstracted [abstractor_datetime] by [abstractor_user]
```

Additional dashboards may be set up to identify passages with unresolved abstractor flags, passages with missing primary outcome, etc.

##### Rule-based auto-daemons

At times, stereotyped records may be amenable to *post hoc* automated adjudication using rule-based strategies or other algorithms. Such strategies may be implemented outside of REDCap and their results imported with a unique value for `coreview_outcome` (in our implementation, 99) to allow further handling by dynamic record pooling APIs (**Appendix D**).
