## Appendix B for "A modular pipeline for natural language processing-screened human abstraction of a pragmatic trial outcome from electronic health records"

### Appendix B. Source code for preparing EHR data for in-context display in REDCap.

Example Stata version 18 (StataCorp, stata.com) source code for preparing EHR data for in-context display in REDCap. Accepts input file with patient identifiers, note identifiers, EHR passage (segment) identifiers, segment text (as UTF-8 plaintext), and numerical NLP-predicted passage-level probabilities. Replaces segment text variable with non-standard HTML displaying each screen-positive EHR passage within its surrounding context (REDCap adds `<br>` tags for each newline).

```
version 18
clear all

// define numerical threshold for NLP positivity
local BERT_THRESHOLD_SOFTMAX = -1

// define file path for CSV file of EHR data
local IMPORT_FILE = "segments_and_predicted_probabilities.csv"
/* expected contents:
   pid           deidentified patient identifier
   note_eventid  deidentified note identifier
   segment_id    deidentified *sequential* within-note EHR passage/segment identifier
   segment_txt   passage/segment EHR text
   segment_prob  NLP-predicted probability of outcome within EHR passage/segment
*/

import delim using "`IMPORT_FILE'", bindquotes(strict) maxquotedrows(0) encoding(UTF-8)

// prepend/append segments with surrounding context + formatting
// (restricted to NLP-positive passages for efficiency)

  // generate filter variable for NLP-positive passages
  gen _f = (segment_prob >= `BERT_THRESHOLD_SOFTMAX')

  sort pid note_eventid segment_id
  // IMPORTANT: assumes that segment_id is unique and sequentially ordered within each note

  local two_newlines="char(10)+char(10)"
  // IMPORTANT: May need to customize -two_newlines- to reflect line endings of local system.
  //            - Stata char(10) = '\n' (LR; line feed) is a newline in MacOS X and UNIX/Linux systems.
  //            - Stata char(13) = '\r' (CR; carriage return)
  //            - Stata char(13)+char(10) (LF+CR) is a newline in Microsoft Windows systems.

  // original segment
```

```
  // - unadorned in formatting; RedCAP appends <br> to newlines
  gen       segment_incontext = segment_txt if _f

  // prepend segments x 1-2 (prepends two if most proximal segment is <200 characters)
  // - <span> tags reformat surrounding context
  replace   segment_incontext = `"<span style="color:#888888;font-size:11pt;">"' + segment_txt[_n-1] + "</span>" + `two_newlines' + segment_incontext   if _f & note_eventid[_n-1]==note_eventid[_n]
  replace   segment_incontext = `"<span style="color:#888888;font-size:11pt;">"' + segment_txt[_n-2] + "</span>" + `two_newlines' + segment_incontext   if _f & note_eventid[_n-2]==note_eventid[_n] & strlen(segment_txt[_n-1])<200

  // append segments x 1-2 (appends two if most proximal segment is <200 characters)
  // - <span> tags reformat surrounding context
  replace   segment_incontext = segment_incontext + `two_newlines' + `"<span style="color:#888888;font-size:11pt;">"' + segment_txt[_n+1] + "</span>"   if _f & note_eventid[_n+1]==note_eventid[_n]
  replace   segment_incontext = segment_incontext + `two_newlines' + `"<span style="color:#888888;font-size:11pt;">"' + segment_txt[_n+2] + "</span>"   if _f & note_eventid[_n+2]==note_eventid[_n] & strlen(segment_txt[_n+1])<200

  // overwrite segment_txt
  drop segment_txt
  rename segment_incontext segment_txt

// restrict to NLP+ passages
keep if _f
```
