## Appendix C for "A modular pipeline for natural language processing-screened human abstraction of a pragmatic trial outcome from electronic health records"

### Appendix C. Source code for temporal-sequence-preserving sort of passage-level EHR data.

Example Stata version 18 (StataCorp, stata.com) source code for performing temporal-sequence-preserving sort of passage- and note-level electronic health record (EHR) data to facilitate human adjudication of time-to-event data.

Users of versions of Stata preceding version 16 may replicate this functionality by using `preserve` and `restore` in lieu of creating a new data frame (herein named `sort_notes_within_patients`), saving the contents of said data frame into a `tempfile` and then using `merge` to merge the original data with the `tempfile`.

```
version 18
clear all

// construct a simulated dataset

  // simulate list of 1,000 patients with randomly ordered patient IDs (patient_id) and numbers of notes
  set obs 1000
  gen patient_id = runiformint(1,10000)
  gen date_start = runiformint(date("1/1/2020","MDY"),date("12/31/2022","MDY"))
  gen notes_per_patient = runiformint(1,50)
  format %td date_start

  // expand to 1 observation per EHR note
  expand notes_per_patient
  gen note_id = _n
  gen note_date = runiformint(date_start, date_start+30)
  gen passages_per_note = runiformint(20,100)
  format %td note_date

  // expand to 1 observation per EHR passage
  expand passages_per_note
  gen passage_id = _n

  // simulate NLP screen positive for 1% of passages, and drop negative passages
  gen nlp_screen = runiform() < .01
  drop if nlp_screen == 0

// ----------------------------------------------------------------------------

// sort dataset in order of patient ID -> ascending note date -> random, then generate within-patient passage identifier
gen _shuffle = runiform()
sort patient_id note_date _shuffle
by patient_id: gen passage_id_withinpatient = _n
```

```
// shuffle dataset, preserving temporal sequence of notes within each patient
sort _shuffle
gen seq = _n
frame put patient_id seq passage_id_withinpatient, into(sort_notes_within_patients)
frame sort_notes_within_patients {
  keep patient_id seq passage_id_withinpatient
  reshape wide seq, i(patient_id) j(passage_id_withinpatient)
  ds seq*
  local nseq: word count `r(varlist)'
  rowsort seq*, gen(sorted_seq1-sorted_seq`nseq')
  drop seq*
  reshape long sorted_seq, i(patient_id) j(passage_id_withinpatient)
  drop if mi(sorted_seq)
  assert sorted_seq[_n] > sorted_seq[_n-1] if patient_id[_n]==patient_id[_n-1]
}
frlink 1:1 patient_id passage_id_withinpatient, frame(sort_notes_within_patients)
frget sorted_seq, from(sort_notes_within_patients)
sort sorted_seq

// error-check result to ensure monotonicity of note_date within patient within shuffled sequence
gsort patient_id note_date _shuf
assert sorted_seq[_n] > sorted_seq[_n-1] if patient_id[_n]==patient_id[_n-1]

// set REDCap record ID to sorted_seq
rename sorted_seq record_id
order record_id, first
sort record_id
```
