## Appendix D for "A modular pipeline for natural language processing-screened human abstraction of a pragmatic trial outcome from electronic health records"

### Appendix D. Dynamic record pooling script for time-to-first-event data collection.

This is a `bash` (Bourne-again shell, v3.2) script that uses the REDCap application programming interface (API) to identify and "demote" electronic health record (EHR) passages that may be discarded from the minimum complete dataset for human adjudication, with the goal of a **data completion for time-to-first-event**. The provided structured query language (SQL) queries may be modified to accommodate other minimum complete dataset definitions. The script may be run by manual invocation, or may be executed by an automated process (e.g., `cron`). In our implementation, we designated a `cron` process to run the script every evening.

**Notes:**

- This script is dependent on the `csvkit` data manipulation library by Christopher Groskopf et al, available under the open-source MIT License at https://csvkit.readthedocs.io/en/latest/; and, on the `curl` tool for secure Hypertext Transfer Protocol (HTTPS) transfers, available under an open-source license at https://curl.se.
- This script assumes that the temporary files it creates via `mktemp` are secure, and that the RedCAP API implementation and HTTPS calls to it are also secure.
- This script retrieves a REDCap *primary outcome report* which must be set up through REDCap. Although the REDCap API itself is secured by a token, further safeguards may be enacted by restricting the REDCap API token to non-protected health information (PHI) fields, and by restricting the contents of the *primary outcome report* to non-PHI fields. Our *primary outcome report* consisted of the fields `record_id`, `pid`, `note_eventid`, `segment_id`, `outcome_human`, `abstractor_flag___1`, `abstractor_flag___2`, and `coreview_outcome`. Values of `pid`, `note_eventid`, and `segment_id` were hashed to remove PHI identifiers prior to importing into REDCap.
- `DYNABS_DEBUG_MODE` and `DYNABS_RENAME_ARRAYS_OPTION` are provided to allow testing and debugging of the script prior to deploying it on actual project data. Values for both may be overridden in the command line using the `-d` and `-a` options documented in the source code.
- The algorithm for determining which records should be discarded from, or replaced into, the minimum complete dataset is implemented in the two invocations of `csvsql`. In this implementation, records with unresolved abstractor flags (i.e., `abstractor_flag___1` = 1) are not considered human-confirmed outcomes.
- Records are discarded from the minimum complete dataset by adding `PREFIX_FOR_SKIP` (default value of $9 \times 10^8$) to the `record_id`, effectively moving the record to the bottom of REDCap's sort order. This places an implicit limit on the number of records that may be handled.
- **IMPORTANT: Records discarded from the minimum complete dataset will still appear in the database, with all extant human adjudications preserved.** This design allows the system to be robust to changes in data (e.g., handling of changing abstraction results following co-review), but requires due consideration for interpretation of exported data.
- To accommodate *post hoc* rule-based auto-daemons that may adjudicate records outside of REDCap,

records with `coreview_outcome` value equaling a discrete value (i.e., 99) are automatically discarded from the minimum complete dataset (see *Rule-based auto-daemons* in **Appendix A**).

```
#!/bin/bash

#
# Dependencies: `csvkit` <https://csvkit.readthedocs.io/en/latest/>
#       `curl` <https://curl.se>
#

# ------------------------------------------------------------------------------

# RedCap API Token and configuration parameters
#
REDCAP_TOKEN="YOUR_API_KEY_HERE"
REDCAP_REPORT_ID="YOUR_PRIMARY_OUTCOME_REPORT_ID_HERE"
REDCAP_API_URL="YOUR_REDCAP_SERVER_API_URL_HERE, e.g., https://redcap.institution/api/"

# Log file location, empty string to suppress
# Note: Logging is disabled in debug modes, as output goes to stdout.
#
DYNABS_LOG_FILE="YOUR_LOGFILE_PATH_HERE/logfile.log"

# Location for backups of RedCap primary outcome report (with trailing slash and
# filename prefix if desired; empty string to suppress; disabled in debug modes
#
DYNABS_REDCAP_REPORT_BACKUPS_PATHPREFIX="YOUR_BACKUP_PATH_HERE/backup-"

# Debug modes:
#   0: Normal mode: pulls primary outcome report from RedCap API, and executes
#      promotions/demotions via RedCap API
#   1: File mode: accepts .csv file for primary outcome report via option -f,
#      then generates promotions/demotions but does not execute in RedCap
#   2: Retrieval mode: pulls primary outcome report from RedCap API, then
#      generates promotions/demotions but does not execute in RedCap
# Logging is disabled in debug modes, as output goes to stdout.
# In debug mode 1, pass .csv file using option -f
#
# Hard-coded default here, may override by command-line options below:
#
DYNABS_DEBUG_MODE=0

# Array option: Controls output of rename arrays (which can be very long) in
# debug modes; no effect when not in a debug mode. Modes:
#   0: suppress
#   1: output readable format
#   2: output untransformed
#
```

```
# Hard-coded default here, may override by command-line options below:
#
DYNABS_RENAME_ARRAYS_OPTION=0

# Command line options for overriding debug mode and array options:
#
# Usage: (command.sh) [-d debug_mode] [-f input_filename] [-a array_option]
#        - debug_mode: 0 normal, 1 file mode, 2 retrieval mode
#        - input_filename: path to .csv file -- required for debug file mode
#        - array_option: 0 suppress, 1 readable format, 2 untransformed format
#
while getopts ":d:f:a:" opt; do
  case ${opt} in
    d )
      DYNABS_DEBUG_MODE="${OPTARG}"
      ;;
    f )
      DYNABS_INPUT_FILENAME="${OPTARG}"
      ;;
    a )
      DYNABS_RENAME_ARRAYS_OPTION="${OPTARG}"
      ;;
    \? )
      echo "Invalid option: -$OPTARG" >&2
      exit 1 ;;
    : )
      echo "Option -$OPTARG requires an argument." >&2
      exit 1 ;;
  esac
done

# Check for valid file argument if debug mode 1
if [[ "$DYNABS_DEBUG_MODE" -eq 1 ]]; then
  if [[ -z "$DYNABS_INPUT_FILENAME" ]]; then
    echo "Error: The -f filename option is required when -d 1 (file mode) is selected." >&2
    exit 1
  elif [[ ! -e "$DYNABS_INPUT_FILENAME" ]]; then
    echo "Error: File '$DYNABS_INPUT_FILENAME' does not exist." >&2
    exit 1
  fi
fi

# Promotion and demotion of records for abstraction takes place by adding or
# subtracting PREFIX_FOR_SKIP to/from the record_id.
#
# Records with record_id > PREFIX_FOR_SKIP are demoted for abstraction.
#
PREFIX_FOR_SKIP=900000000
```

```
if [[ $PREFIX_FOR_SKIP -ge 2147483647 ]]; then
  echo "Error: PREFIX_FOR_SKIP exceeds range of 32-bit signed integers." > /dev/stderr
  exit 1
fi

# --------------------------------------------------------------------------------

# Determine output of logging (debug modes -> stdout; blank log file -> null)
# Initiates log file if indicated
if [ $DYNABS_DEBUG_MODE -gt 0 ]; then
  DYNABS_LOG_DESTINATION="/dev/stdout"
elif [[ -z "$DYNABS_LOG_FILE" ]]; then
  # empty string for DYNABS_LOG_FILE
  DYNABS_LOG_DESTINATION="/dev/null"
else
  DYNABS_LOG_DESTINATION="$DYNABS_LOG_FILE"
fi
echo >> $DYNABS_LOG_DESTINATION
echo >> $DYNABS_LOG_DESTINATION
echo "--- $(date) ---" >> $DYNABS_LOG_DESTINATION

# Get primary outcome report from RedCap API
REDCAP_CSV=$(mktemp)
if [ $DYNABS_DEBUG_MODE -eq 1 ]; then
  REDCAP_CSV_ORIG="$DYNABS_INPUT_FILENAME"
  if [[ ! -e "$REDCAP_CSV_ORIG" ]]; then
    echo "Error: You are in debug mode 1. Please specify a valid .csv file using option -f." > /dev/stderr
    exit 1
  fi
else
  if [[ ! -z "$DYNABS_REDCAP_REPORT_BACKUPS_PATHPREFIX" ]]; then
    REDCAP_CSV_ORIG="$DYNABS_REDCAP_REPORT_BACKUPS_PATHPREFIX$(date +"%Y%m%d-%H%M%S").csv"
  else
    REDCAP_CSV_ORIG=$(mktemp)
  fi
  DATA="token=$REDCAP_TOKEN&content=report&format=csv&report_id=$REDCAP_REPORT_ID&csvDelimiter=&rawOrLabel=raw&rawOrLabelHeaders=raw&exportCheckboxLabel=false&returnFormat=json"
  CURL=`which curl`
  $CURL -H "Content-Type: application/x-www-form-urlencoded" \
        -H "Accept: application/json" \
        -X POST \
        -d "$DATA" \
        $REDCAP_API_URL \
        > "$REDCAP_CSV_ORIG"
fi
```

```
# ---------------------------------------------------------------------------

# Calculate record_id_orig, and add it as a column to the .csv file
awk -F',' -v OFS=',' -v prefixForSkip=$PREFIX_FOR_SKIP '
NR == 1 {
  header = $0
  print header ",record_id_orig"  # Print original header plus new column
  next
}
{
  record_id = $1
  if (record_id > prefixForSkip) {
    record_id_orig = record_id - prefixForSkip
  } else {
    record_id_orig = record_id
  }
  print $0, record_id_orig
}' "$REDCAP_CSV_ORIG" > "$REDCAP_CSV"

# ---------------------------------------------------------------------------

# Initialize arrays for rename requests
RECS_RENAME_OLDNAMES=()
RECS_RENAME_NEWNAMES=()

# Identify non-demoted records with positive outcome and negative flag, and
#   generate arrays of subsequent records to demote
# Also demote anything with coreview_outcome==99 (i.e. Dupinator or other
#   rule-based daemons) that hasn't already been demoted
RECS_TO_DEMOTE=$(csvsql --tables redcap --query "
  WITH verifiedPositives AS (SELECT * FROM redcap WHERE outcome_human = 1 AND abstractor_flag___1 = 0)
  SELECT DISTINCT CAST (redcap.record_id AS INTEGER) AS record_id
  FROM verifiedPositives JOIN redcap ON verifiedPositives.pid = redcap.pid
  WHERE (redcap.coreview_outcome = 99 AND redcap.record_id < $PREFIX_FOR_SKIP)
    OR
  (verifiedPositives.record_id_orig < redcap.record_id_orig
    AND redcap.record_id < $PREFIX_FOR_SKIP
    AND redcap.outcome_human IS NULL)
" "$REDCAP_CSV" | tail -n +2)

if [[ -n "$RECS_TO_DEMOTE" ]]; then
  while IFS= read -r REC_ID; do
    RECS_RENAME_OLDNAMES+=("$REC_ID")
    RECS_RENAME_NEWNAMES+=("$((REC_ID + PREFIX_FOR_SKIP))")
  done <<< "$RECS_TO_DEMOTE"
fi

```

```
# Output debugging code
  echo >> $DYNABS_LOG_DESTINATION
  echo -n "Records to be demoted: " >> $DYNABS_LOG_DESTINATION
  echo "$RECS_TO_DEMOTE" | sort -n | tr '\n' ' ' >> $DYNABS_LOG_DESTINATION
  echo >> $DYNABS_LOG_DESTINATION
  echo -n "Number to be demoted: " >> $DYNABS_LOG_DESTINATION
  echo "$RECS_TO_DEMOTE" | wc -w | xargs >> $DYNABS_LOG_DESTINATION
  echo >> $DYNABS_LOG_DESTINATION

# ------------------------------------------------------------------------------

# Identify previously-demoted records that do NOT (i.e., no longer) follow a
#   previous unflagged confirmed-positive, and generate arrays of records to
#   promote
# * excludes anything with coreview_outcome==99 (i.e. Dupinator or other
#   rule-based daemons)
RECS_TO_PROMOTE=$(csvsql --tables redcap --query "
  WITH verifiedPositives AS (SELECT * FROM redcap WHERE outcome_human = 1 AND abstractor_flag___1 = 0),
  demoted AS (SELECT * FROM redcap WHERE record_id > $PREFIX_FOR_SKIP AND (coreview_outcome != 99 OR coreview_outcome IS NULL))
  SELECT DISTINCT CAST (demoted.record_id AS INTEGER) AS record_id
  FROM demoted LEFT JOIN verifiedPositives ON demoted.pid = verifiedPositives.pid AND verifiedPositives.record_id_orig < demoted.record_id_orig
  WHERE verifiedPositives.record_id IS NULL AND (demoted.outcome_human IS NULL OR demoted.abstractor_flag___1 = 1)
" "$REDCAP_CSV" | tail -n +2)

if [[ -n "$RECS_TO_PROMOTE" ]]; then
  while IFS= read -r REC_ID; do
    RECS_RENAME_OLDNAMES+=("$REC_ID")
    RECS_RENAME_NEWNAMES+=("$((REC_ID - PREFIX_FOR_SKIP))")
  done <<< "$RECS_TO_PROMOTE"
fi

# Output debugging code
  echo >> $DYNABS_LOG_DESTINATION
  echo -n "Records to be promoted: " >> $DYNABS_LOG_DESTINATION
  echo "$RECS_TO_PROMOTE" | sort -n | tr '\n' ' ' >> $DYNABS_LOG_DESTINATION
  echo >> $DYNABS_LOG_DESTINATION
  echo -n "Number to be promoted: " >> $DYNABS_LOG_DESTINATION
  echo "$RECS_TO_PROMOTE" | wc -w | xargs >> $DYNABS_LOG_DESTINATION
  echo >> $DYNABS_LOG_DESTINATION

# ------------------------------------------------------------------------------

# Send rename requests to RedCap API (only supports sending them one at a time)
if [ $DYNABS_DEBUG_MODE -eq 0 ]; then
  echo "curl output:" >> $DYNABS_LOG_DESTINATION
```

```
  CURL=`which curl`
  for ((i=0; i<${#RECS_RENAME_OLDNAMES[@]}; i++)); do
    echo "Record $((i+1)) of ${#RECS_RENAME_OLDNAMES[@]}:"
    echo -n "  rename ${RECS_RENAME_OLDNAMES[$i]} -> ${RECS_RENAME_NEWNAMES[$i]}: " >> $DYNABS_LOG_DESTINATION
    DATA="token=$REDCAP_TOKEN&action=rename&content=record&record=${RECS_RENAME_OLDNAMES[$i]}&new_record_name=${RECS_RENAME_NEWNAMES[$i]}&returnFormat=json"
    $CURL -H "Content-Type: application/x-www-form-urlencoded" \
          -H "Accept: application/json" \
          -X POST \
          -d "$DATA" \
          $REDCAP_API_URL \
          >> $DYNABS_LOG_DESTINATION
    echo >> $DYNABS_LOG_DESTINATION
    sleep 2      # added to honor institutional API call limits
  done
fi

# Output debugging code for rename arrays in debug mode only; never logged
if [[ $DYNABS_DEBUG_MODE -gt 0 && $DYNABS_RENAME_ARRAYS_OPTION -gt 0 ]]; then
  if [ $DYNABS_RENAME_ARRAYS_OPTION -eq 1 ]; then
    echo
    echo "Renaming arrays (readable): "
    for ((i=0; i<${#RECS_RENAME_OLDNAMES[@]}; i++)); do
      printf "${RECS_RENAME_OLDNAMES[$i]}\t->\t${RECS_RENAME_NEWNAMES[$i]}\n"
    done
    echo
  else
    echo
    echo "Renaming arrays (raw): "
    echo "RECS_RENAME_OLDNAMES: ${RECS_RENAME_OLDNAMES[@]}"
    echo "RECS_RENAME_NEWNAMES: ${RECS_RENAME_NEWNAMES[@]}"
    echo
  fi
fi
```
